# Adverse effects of COVID-19 related lockdown on pain, physical activity and psychological wellbeing in people with chronic pain

**DOI:** 10.1101/2020.06.04.20122564

**Authors:** Nicholas Fallon, Christopher Brown, Hannah Twiddy, Eleanor Brian, Bernhard Frank, Turo Nurmikko, Andrej Stancak

## Abstract

Countries across the world imposed lockdown restrictions during the COVID-19 pandemic. It has been proposed that lockdown conditions disproportionately impact those living with chronic pain, requiring adaptation to treatment and care strategies. We investigated how lockdown restrictions in the United Kingdom impacted individuals with chronic pain (N = 431) relative to a healthy control group (N = 88) using an online survey. In accordance with the fear-avoidance model, we hypothesised increases in perceived pain and psychological distress that would be mediated by pain catastrophizing. Survey questions answered during the lockdown period, probing patients’ self-perceived changes retrospectively, revealed that people with chronic pain perceived increases in their pain severity compared to before lockdown. They were also more adversely affected by lockdown compared to pain-free individuals, demonstrating greater increases in anxiety and depressed mood, increased loneliness and reduced levels of physical exercise. Pain catastrophizing was found to be an important factor in predicting the extent of self-perceived increases in pain, and accounted for the relationship between decreased mood and pain. Perceived decreases in levels of physical exercise also independently predicted perceptions of increased pain. Interestingly, actual changes in pain symptoms (measured at two time points at pre- and post-lockdown in a subgroup, N = 85) did not change significantly on average, but those reporting increases also demonstrated greater baseline levels of pain catastrophizing. Overall, the findings suggest that remote pain management provision to target reduction of catastrophizing and increases to physical activity could be beneficial for chronic pain patients in overcoming the adverse effects of lockdown.

## 1. Introduction

COVID-19 is a highly contagious disease related to the spread of SARS-CoV-2 virus [17]. Due to the high infection and mortality rate of COVID-19, many countries implemented periods of lockdown to reduce uncontrolled spread of the virus [25]. Lockdown of economic and social activities creates a situation of threat in vulnerable populations due to health anxiety, physical inactivity, reduced accessibility to usual care, social isolation, and financial-economic uncertainty [25; 42].

It was recently proposed that the COVID-19 pandemic would substantially impact those living with chronic pain, and thus require efforts to adapt treatment and care strategies [21]. Chronic pain affects around 40% of the UK adult population [23], and represents a significant global burden at both the individual and socioeconomic level [8; 43]. Increased prevalence of chronic pain in the elderly and those with comorbid illness or disability [20; 23] overlaps with the highest risk for COVID-19. Empirical research is essential to capture how people living with chronic pain are affected by the current pandemic, and to support efforts to develop pain management approaches in these challenging conditions, e.g., online technologies to improve levels of social support, combat social-isolation and offer treatment provision [4; 37].

Previous research indicates a likelihood that chronic pain populations suffer increased severity of symptoms in high-stress situations including war or the aftermath of terrorist attacks [14; 35]. If we can better understand how high-stress situations exacerbate chronic pain, we can adapt clinical strategies to mitigate the associated suffering [47]. A likely mediator of greater pain severity resulting from high-stress situations is psychological distress [33], which critically impacts on the perception of pain, physical disability [22], and overall quality of life [7; 11; 28]. For example, anxiety augments neural processes modulating the perception of pain [18; 41; 57]. In addition, the fear-avoidance model of chronic pain [29; 55] points to the theoretical importance of pain-related fear and catastrophizing as contributors to decreased mood and physical activity, which in turn exacerbate pain symptoms. A unique characteristic of the COVID-19 lockdowns are physical and social distancing measures; these measures would be expected to impact pain symptoms via a combination of changes in physical activity levels, mood and anxiety. Reduced physical activity during the COVID-19 pandemic could exacerbate effects of psychological stress and reduce coping with anxiety and depression, especially in vulnerable populations [27].

The impact of COVID-19 on mental health is becoming increasingly apparent. Increased psychological distress is evident in COVID-19 patients and health professionals who treat them [12; 31]. During peak lockdown conditions, sharp increases were seen in prevalence of anxiety and depression in the general adult population in China [30]. In research from the United States and Spain, ‘stay at home’ directives and living with chronic illness are factors associated with greater risk of adverse effects [39; 53]. Considering this evidence, there is a clear need to understand how changes in psychological wellbeing and physical activity levels, due to the ongoing pandemic and related lockdown conditions, impact on pain experience in chronic pain populations.

## 2. Method

### 2.1. Aims and hypotheses

The present study aimed to capture the effects of the COVID-19 pandemic, and corresponding UK lockdown restrictions, on pain, psychological wellbeing and physical activity levels in a group of participants suffering from chronic pain compared to a non-pain group. We hypothesized that lockdown conditions would cause increased levels of pain severity relative to pre-lockdown period in those with chronic pain. Secondly, we predicted that lockdown conditions would have a greater impact on the psychological and physical wellbeing of chronic pain, relative to non-pain, respondents. Thirdly, we hypothesised that self-perceived changes in reported pain levels could be due to levels of pain catastrophizing, and changes in their psychological wellbeing and physical activity in accordance with theoretical models of fear-avoidance.

### 2.2. Design and procedure

Participants (N = 519) took part in an online design comprising self-reporting chronic pain participants (N = 431) and a comparison sample of non-pain control participants (N = 88). All participants were recruited via targeted online advertisements to relevant forums or support groups, or via direct invitation to chronic pain patients who had previously given the experimenters permission to be contacted for future research. The latter option engendered a subgroup from whom baseline data on pain and psychological measures were available (N = 85). Responding participants were directed to the study pages which were programmed in Qualtrics software (Qualtrics, Utah, USA). Firstly, participants read an information sheet and gave informed consent using a tick box procedure. They answered demographic questions, self-reported whether they had chronic pain and their relevant diagnosis, and answered some questions about their personal lockdown conditions such as size of household. A series of visual analogue scales captured current pain and wellbeing levels before participants completed self-report differential measures indicating their self-perception of change in their pain, physical exercise and wellbeing relative to pre-COVID levels. Finally, participants completed a series of short, validated questionnaires to capture pain, pain related cognition and psychological wellbeing (full description below). A debrief page at the end of the study provided information on the purpose of the study. They were informed of how to contact the researchers directly with any questions and we also highlighted some useful resources for those suffering pain or psychological distress during lockdown. All respondents were reimbursed £3.33 for completing the session as part of an ongoing longitudinal data collection.

### 2.3. Participants and lockdown conditions

Participants (N = 519) took part. This total comprised 470 females, 45 males, and 4 participants who selected ‘other’. Ages ranged from 18 to 79 years (43.98 ± 13.38, mean ± SD). Chronic pain respondents (N = 431) comprised a range of chronic pain conditions. The primary diagnosis was categorized according to the Classification of Diseases (ICD-11) of the World health Organization, derived from the main cause of their pain (Table 1). When a specific diagnosis indicated by the patient it was recorded as a secondary diagnosis, shown as codes only in Table 1. A subgroup of chronic pain respondents (N = 85) were recruited via contacts with a local tertiary care pain clinic, having previously given agreement to be contacted for research purposes. In this subgroup, baseline data on pain and psychological measures was available, which had been collected in the 6 months preceding UK lockdown. Finally, a sample of age and sex matched non-pain control respondents (N = 88) were also collected via online advertisements.

All participants were based in the United Kingdom. First responses were recorded 3.5 weeks after the initiation of UK lockdown conditions on April 17^th^ 2020, final responses were recorded on May 12^th^ 2020. This period covered the most stringent level of lockdown in UK, comprising social distancing and advice against all non-essential travel with recommendations to work from home. Exercise with social distancing was permitted once per day. Enhanced lockdown recommendations were in place for those deemed high-risk [1]. UK recommendations were relaxed on May 13^th^ 2020 and the data collection was halted.

**Table 1.**
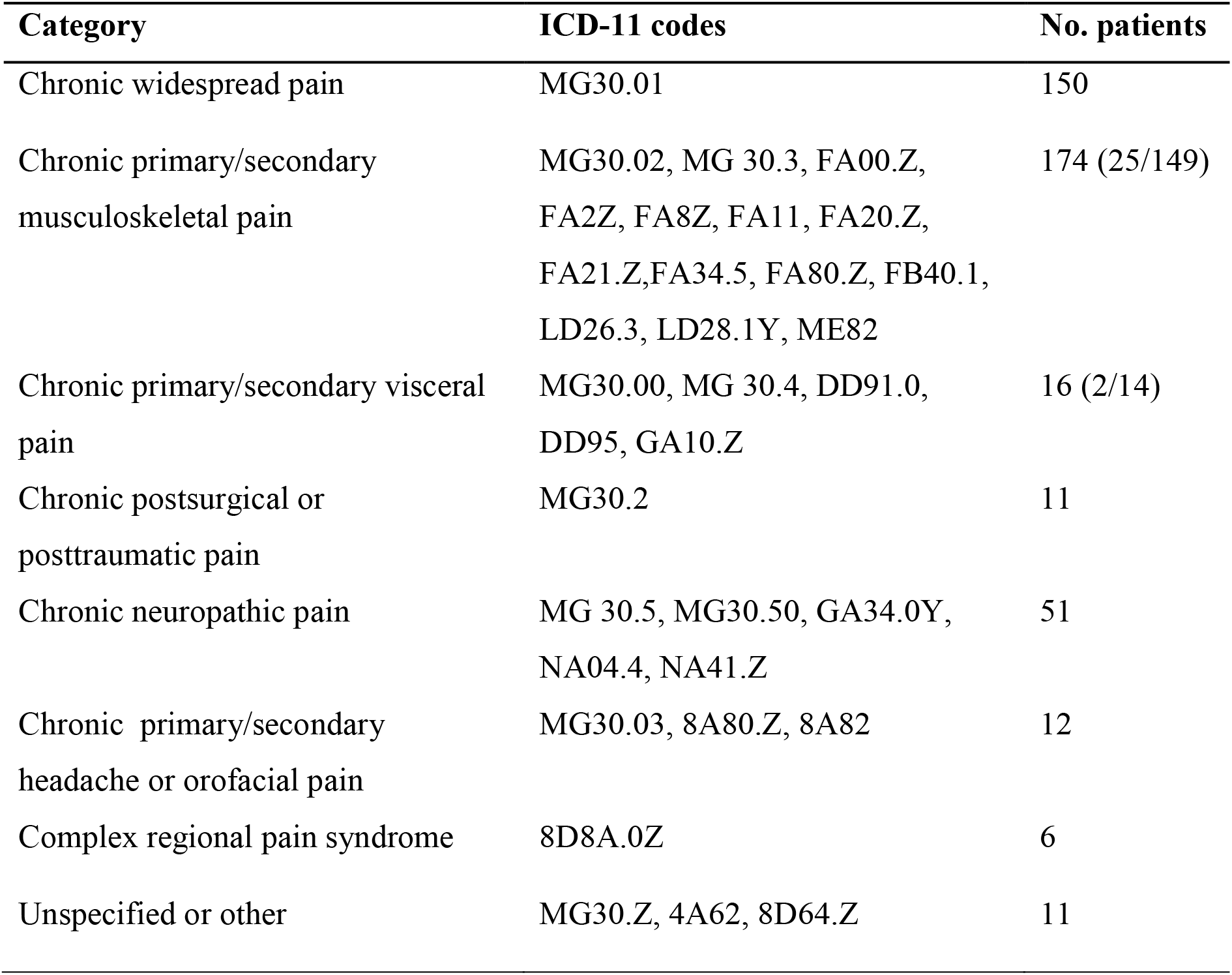
Number of patients corresponding to each diagnostic category, and disease identification code, according to ICD-11 guidelines.

| Category | ICD-11 codes | No. patients |
| --- | --- | --- |
| Chronic widespread pain | MG30.01 | 150 |
| Chronic primary/secondary musculoskeletal pain | MG30.02, MG 30.3, FA00.Z, FA2Z, FA8Z, FA11, FA20.Z, FA21.Z,FA34.5, FA80.Z, FB40.1, LD26.3, LD28.1Y, ME82 | 174 (25/149) |
| Chronic primary/secondary visceral pain | MG30.00, MG 30.4, DD91.0, DD95, GA10.Z | 16 (2/14) |
| Chronic postsurgical or posttraumatic pain | MG30.2 | 11 |
| Chronic neuropathic pain | MG 30.5, MG30.50, GA34.0Y, NA04.4, NA41.Z | 51 |
| Chronic primary/secondary headache or orofacial pain | MG30.03, 8A80.Z, 8A82 | 12 |
| Complex regional pain syndrome | 8D8A.0Z | 6 |
| Unspecified or other | MG30.Z, 4A62, 8D64.Z | 11 |

### 2.4. Self-report measures

Participants were asked whether they currently suffered from chronic pain. Those who answered affirmatively completed follow-up questions about the intensity of their pain in the previous week using a visual analogue scales (0–100, anchors “No pain at all” to “Extremely Severe Pain”). All participants completed further VAS scales for described levels of the following variables for the previous week: tiredness (0–100, anchors “Not at all” to “Extremely tired”); loneliness (0–100, anchors “Not at all” to “Extremely lonely”); and anxiety (0–100, anchors “Not at all” to “Extremely anxious”).

Participants who reported chronic pain then completed a differential scale, where they rated perception of pain intensity in the past 7 days relative to a typical week in the pre-COVID period. Again, this utilised a VAS (0–100, anchors “Very much better”, centre marker “About the same”, to “Very much worse”). All participants completed differential VAS scales to indicate their perceived change (relative to a typical week in pre-COVID period) for the levels of mood, anxiety and exercise over the past 7 days. The items specifically asked “How tense, nervous or anxious have you felt? (0–100, anchors “Very much better”, centre marker “About the same”, to “Very much worse”), how depressed or blue have you felt?” (0–100, anchors “Very much better”, centre marker “About the same”, to “Very much worse”), “how much physical exercise have you managed to take?” (0–100, anchors “Very much more than usual”, centre marker “About the same”, to “Very much less than usual”). The wording for the questions and anchors for VAS scales differential items was adapted from similar items in the Fibromyalgia Impact Questionnaire [10].

Participants in the chronic pain group then reported any ‘difficulties obtaining pain medication, other treatments or social care in the past two weeks’. All participants were asked whether they had experienced any illness other than chronic pain in the previous two weeks. They also reported whether they were self-isolating due to high-risk status, which encompassed following enhanced recommendations to completely shield oneself during lockdown [1].

Finally, participants completed a series of brief, validated questionnaires to consider pain experience, pain cognition and psychological wellbeing. Specifically, these included the Pain Catastrophizing Scale [PCS, 51], an adapted version of the Brief pain Inventory (BPI) short form [15] and the Hospital Anxiety and Depression Scale [HADS, 58]. The adaptations to the BPI included removal of items requesting patients draw pain location, as well as items on minimal pain and medication lists. These changes were included to optimise the survey for online delivery and reduce the overall time requirement for patients.

### 2.5. Ethics and data sharing

The study was conducted in line with the recommendations of the Declaration of Helsinki and was approved by the local University of Liverpool Research Ethics Committee. The data that support the findings of this study are openly available here doi.10.6084/m9.figshare.12424661.

## 3. Results

### 3.1. Data reduction

A total of 933 participants accessed the study. 135 failed the pre-screening questions which aligned to the exclusion criteria (requiring participants to be > 18 years old and resident in UK during the pandemic). A further 21 did not provide consent after reading the information sheet. There were 65 respondents who consented to take part but did not complete a single item and a further 193 began the study but abandoned without completing a suitable amount of the items to be considered for inclusion (< 90%).

### 3.2 Effect of lockdown on pain intensity, psychological distress and physical activity

Pain was measured in chronic pain respondents by evaluating their perception of average pain intensity for the past week using a 100 point VAS, and also by reporting the differential on their pain intensity relative to a typical week in the pre-lockdown period. The mean pain intensity score in the chronic pain group was 66.64 ± 17.93 (mean ± SD). Univariate t-test analysis indicated that chronic pain respondents reported a statistically significant increase in their pain relative to pre-COVID period on the differential VAS (P < 0.001). The differential scores were numerically transformed to give a score from –100, with negative integers indicating pain decrease, to +100, with positive integers indicative of pain increase (0 values were equal to no perceived change). The mean change for the chronic pain group was 33.64 ± 37.20 (mean ± SD), indicating a significant self-perceived increase in pain compared to the period before the pandemic; t(431) = 18.79, p < 0.001.

To further investigate potential changes in pain intensity relative to pre-COVID periods, a pre-post lockdown comparison was conducted for the subgroup of 85 participants for whom baseline data was available. This group had agreed to take part having already provided previous data in the pre-COVID period (in the 6 months prior to lockdown in the UK). In this subgroup of participants, a within subjects t-test was utilised to compare current pain intensity with previous data. For comparison with existing baseline numerical rating scale (NRS) data, the current pain score was converted from the 100 point VAS to a 10 point NRS equivalent by dividing by 10 and then rounding to nearest whole integer. Five participants had some missing data for the comparison of pain scores and were omitted from the pain comparison. To consider whether changes in psychological pain constructs might also account for different perception of pain levels, we also compared pre-and-post PCS scores (N = 85). Table 2 illustrates mean pain intensity ratings and PCS scores for each time point. Results indicate that patients in the subgroup did not demonstrate a significant increase in reported pain intensity levels compared to data given during the baseline period; t(79) = –1.45, p = 0.15. Likewise there was no significant difference in pain catastrophizing levels captured in the lockdown, relative to baseline, period; t(84) = –0.54, p = 0.59. However, the mean self-perceived change in pain levels for the baseline group was 34.26 ± 33.26 (mean ± SD), indicating a significant self-perceived increase; t(82) = 36.76, p < 0.001 which was comparable to that seen in the full pain cohort ; t(413) = 0.14, p = 0.82.

**Table 2.**
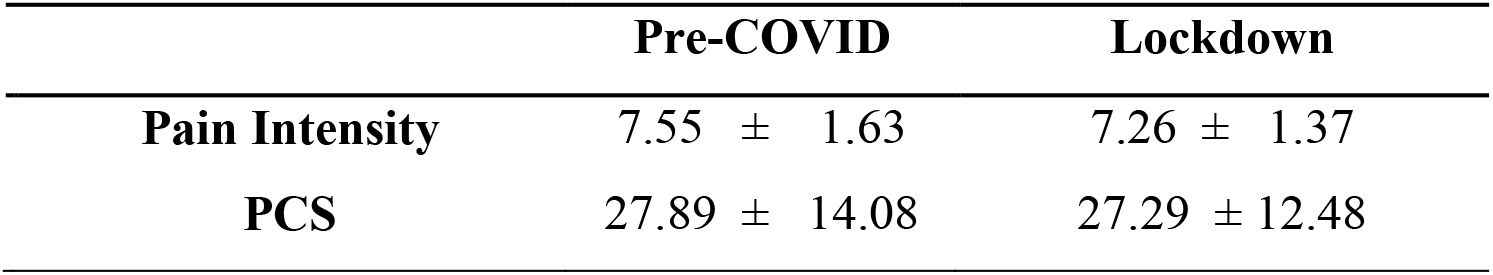
Mean pain intensity (± SD) and pain catastrophizing scores in 85 chronic pain patients for whom the baseline, pre-lockdown data were available.

To further investigate the nature of self-reported changes in pain levels in the subgroup during lockdown, we analysed the relationship between self-reported pain intensity changes and baseline levels of pain catastrophizing recorded prior to the COVID-19 pandemic using Pearson’s correlation analyses. Baseline PCS scores demonstrated a significant correlation with self-reported perception of change in pain levels relative to lockdown periods; r(83) = 0.33, p = 0.003. This suggests that, although direct patient’s reported pain ratings in lockdown may not deviate significantly from those recorded prior to the pandemic, their perception of pain increase in a fashion that aligns to individual differences in pre-existing pain catastrophizing. A comparison of demographic, pain and psychological wellbeing data for the baseline pain group and pain respondents without baseline can be seen in Appendix 1.

### 3.3. Effect of lockdown on chronic pain patients relative to non-pain participants

We hypothesised that participants with chronic pain would demonstrate greater adverse effects of lockdown conditions, indexed by reporting of perceived increases in anxiety and depression, decreases in exercise (relative to the pre-Lockdown periods), and increased scores for loneliness and tiredness. Independent samples t-tests (or Welch’s tests if the assumption of equality of variance was not met) were performed to compare mean ratings across all measures for chronic pain and non-pain groups. Bootstrapping (2000 samples) was used to estimate significance values while mitigating the likelihood of Type I error due to multiple tests. Results indicate that there were no differences between groups on demographics including age and the split of gender. For all variables, the chronic pain group reported significantly greater adverse effects, relative to non-pain participants. Figure 1 and Table 3 illustrate mean scores and comparison statistics for each group.

**Figure 1.**
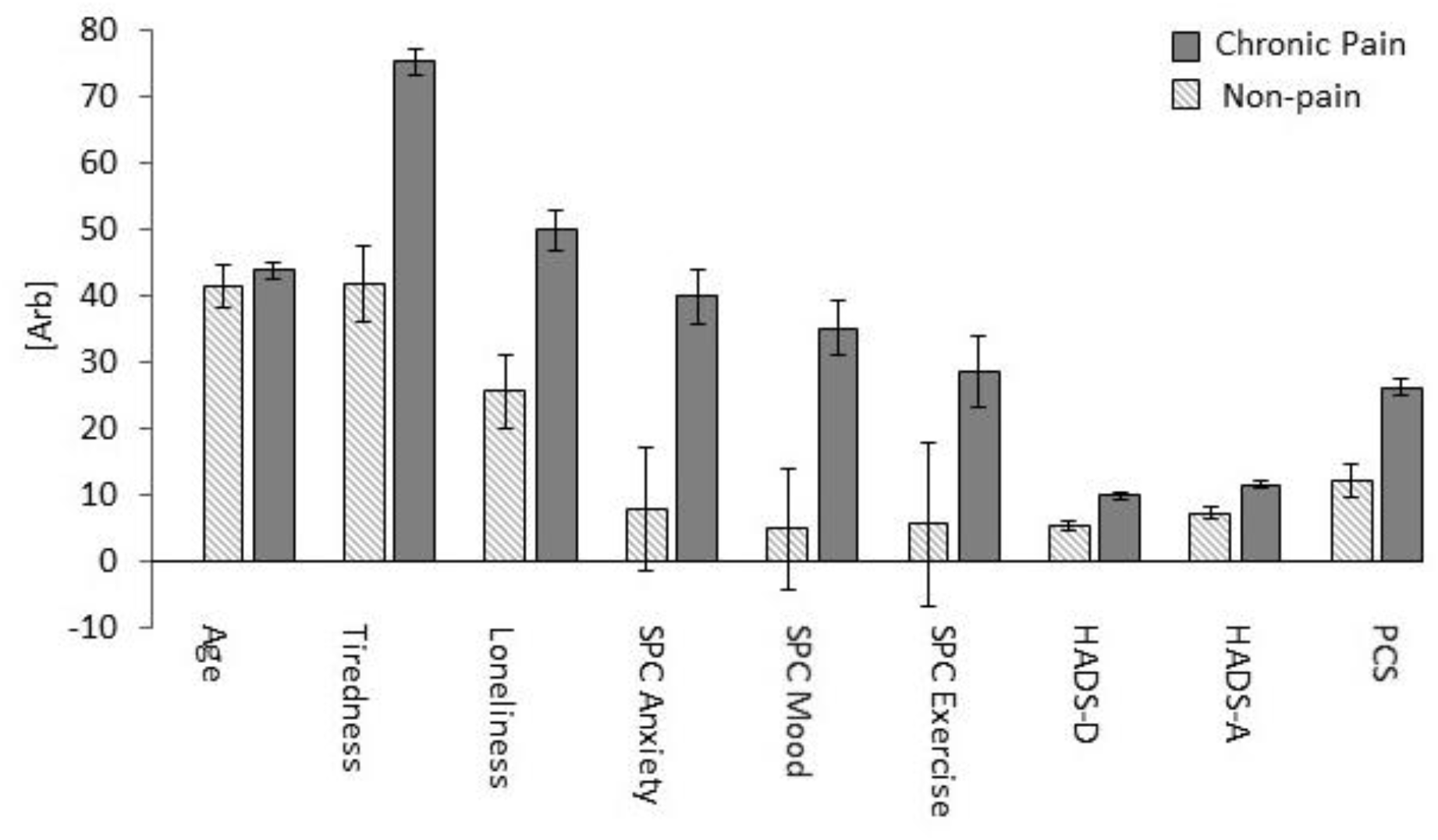
Mean self-reported levels of tiredness and loneliness, self-perceived lockdown-related increases in anxiety, depressed mood and reduction in exercise, HADS-A (anxiety) and HADS-D (depression) and pain catastrophizing (PCS) scores in chronic pain and non-pain respondent groups with standard errors.

**Table 3.** Demographic parameters, self-reported levels of tiredness and loneliness, self-perceived changes (SPC) demonstrating lockdown-related increases in anxiety, depressed mood and reductions in exercise, HADS-A (anxiety) and HADS-D (depression) and pain catastrophizing (PCS) scores in chronic pain and non-pain respondent groups. For each observed measure, means and standard deviations as well as group comparisons using t-test (or Welch’s test) are given with bootstrapped (2000 samples) significance values.

|  | **Chronic pain** | **Non-pain** | **t** | **df** | **P** |
| --- | --- | --- | --- | --- | --- |
| **Sex** | 90.7% Female | 88.7% Female | -.54 | 513 | .55 |
| **Age** | 43.94 ± 13.01 | 41.21 ± 14.98 | 1.59 | 113.87 | .10 |
| **Self-isolating** | 39.68% | 6.81% | 9.26 | 239.66 | <.001 |
| **Any other illness** | 28.07% | 6.81% | 4.63 | 162.60 | <.001 |
| **Anxiety SPC** | 39.36 ± 42.07 | 7.20 ± 42.24 | 6.53 | 517 | <.001 |
| **Depression SPC** | 34.65 ± 41.39 | 4.11 ± 41.52 | 6.30 | 517 | <.001 |
| **Exercise reduction SPC** | 28.69 ± 56.04 | 3.84 ± 57.39 | 3.77 | 512 | .001 |
| **Tiredness** | 75.26 ± 21.03 | 41.66 ± 26.33 | 11.26 | 110.77 | <.001 |
| **Loneliness** | 49.87 ± | 25.43 ± 24.45 | 8.16 | 148.40 | <.001 |
| **HADS-A** | 11.56 ± 4.42 | 7.37 ± 3.57 | 9.56 | 145.33 | <.001 |
| **HADS-D** | 9.86 ± 4.34 | 5.37 ± 3.44 | 9.09 | 512 | <.001 |
| **PCS** | 25.98 ± 12.76 | 12.26 ± 11.34 | 9.21 | 506 | <.001 |

Chronic pain respondents self-reported greater lockdown-related increases in anxiety and depressed mood compared to non-pain group. They also report significant reductions in amount of exercise compared to pre-COVID period whereas negligible reduction was evident in the non-pain group. Chronic pain respondents scored higher on loneliness and tiredness ratings for past 7 days than the non-pain group. Unsurprisingly, increased HADS depression and anxiety scores and increased PCS scores were evident in the chronic pain, relative to non-pain respondents. They also reported increased levels of any other illness in prior 2 weeks (other than chronic pain) compared to non-pain group, and they were more likely to be completely self-isolating due to high risk status.

### 3.4. Self-perceived changes in wellbeing and physical activity predict the self-perceived increase in pain during lockdown for chronic pain participants

We hypothesised that variance in levels of self-reported changes in psychological wellbeing and exercise would predict the degree of perceived increases in pain levels in our chronic pain population. Hierarchical multiple regression analysis was performed to investigate whether self-reported changes in anxiety, depressed mood and physical activity would predict levels of self-reported changes in pain intensity, after controlling for participant age, sex and reports of other illness in the past 2 weeks. Preliminary analyses were conducted to ensure no violation of the assumptions of normality, linearity, and homoscedasticity. In Step 1 of the model, the three confound variables were entered: participant age, sex and reports of other illness. This model was not statistically significant F (3, 415) = 0.43, p = 0.73 and explained 0.5% of variance in self-reported change in pain levels (Table 4). Following entry of self-reported changes in anxiety, depressed mood and exercise in Step 2, the total variance explained by the model was 11% (F (6, 415) = 8. 87; p < 0.001). The introduction of the predictor variables explained an additional 11% of variance in self-reported changes in pain, after controlling for participant age, sex and reports of other illness (R^2^ Change = 0.11; F (3, 415) = 16.96; p < .001). In the final adjusted model, two out of three predictor variables were statistically significant. Self-reported changes in exercise recorded the highest Beta value (β = .17, p < 0.001) followed by changes in depressed mood (β = 0.17, p = 0.008). Changes in anxiety levels were a non-significant predictor (β = 0.11, p = 0.078).

**Table 4.**
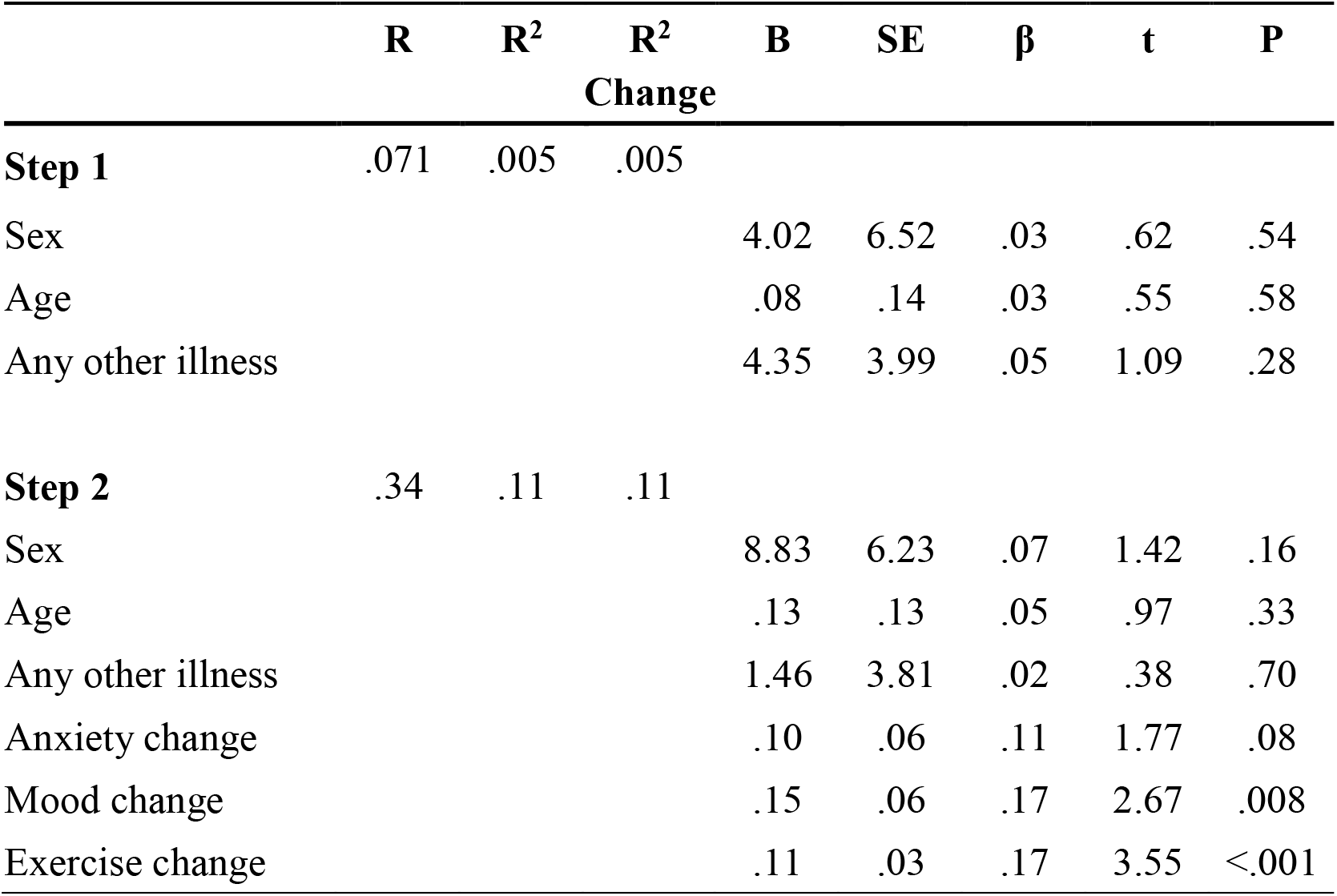
Hierarchical Regression Model of self-reported change in pain intensity. Step 1 describes the inclusion of confound variables prior to the analysis of predictor variables in Step 2. R^2^ = variance explained by IVs; R^2^ Change = additional variance in DV; B = Unstandardized coefficient; β = Standardized coefficient; SE = Standard Error; t = estimated coefficient; P = significance value.

### 3.5. Pain catastrophizing predicts the self-perceived increase in pain during lockdown and mediates the impact of depressed mood in chronic pain participants

In the subgroup of patients with baseline scores, PCS scores from the baseline period before the pandemic demonstrated a significant correlation with perceived levels of change in pain. To investigate whether pain catastrophizing could act as a predictor, and/or mediator, of the relationship between self-perceived changes in mood and exercise and perceived levels of change in pain during lockdown, a mediation regression analysis was performed. The prior hierarchical multiple regression model was repeated with the addition of an intermediary step. After controlling for the confound variables, PCS scores were entered as a mediating variable, before the differential predictors were finally entered. As before, participant age, sex and reports of other illness were entered in Step 1 of the model which was not statistically significant F (3, 415) = 0.67, p = 0.55 (Table 5). Following entry of PCS scores in Step 2 the total variance explained by the model was 12% (F (4, 414) = 13.57; p < 0.001). PCS scores accounted for an additional 11% of variance in self-reported changes in pain, after controlling for participant demographics (R^2^ Change = 0.11; F (1, 411) = 51.96; p < .001). After the inclusion of self-reported changes in mood, anxiety and exercise levels in Step 3 of the model, the total variance explained was 18% (F (4, 411) = 12.63; p < 0.001). The introduction of the differential score predictor variables explained an additional 6% of variance in self-reported changes in pain, after controlling for confounds and PCS scores in the Step 2 (R^2^ Change = 0.06; F (3, 411) = 10.17; p < .001). In the final adjusted model PCS scores exhibited the best predictive value (β = 0.27, p < 0.001), then self-reported changes in physical exercise (β = 0.15, p < 0.001). Change in mood was no longer significant (β = 0.11, p = 0.08), nor were perceived changes in anxiety levels t (β = 0.09, p = 0.17). The analysis indicates that PCS scores are also a significant predictor of self-perceived changes in pain levels during lockdown period. Pain catastrophizing also acts as a partial mediator, as PCS scores accounted for the previously significant relationship between perceived changes in mood and pain, but not for predictive value of perceived changes in exercise.

**Table 5.** Hierarchical Regression Model of self-reported change in pain intensity. R2 = amount of variance explained by IVs; R2 Change = additional variance in DV; B = Unstandardized coefficient; β = Standardized coefficient; SE = Standard Error; t = estimated coefficient; P = significance value.

|  | **R** | **R<sup>2</sup>** | **R<sup>2</sup>**<br>**Change** | **B** | **SE** | **β** | **t** | **P** |
| --- | --- | --- | --- | --- | --- | --- | --- | --- |
| **Step 1** | .071 | .005 | .005 |  |  |  |  |  |
| Sex |  |  |  | 4.02 | 6.55 | 0.03 | 0.61 | 0.54 |
| Age |  |  |  | 0.08 | 0.14 | 0.03 | 0.55 | 0.58 |
| Any other illness |  |  |  | 4.35 | 4.01 | 0.05 | 1.09 | 0.28 |
| **Step 2** | .34 | .11 | .11 |  |  |  |  |  |
| Sex |  |  |  | 1.90 | 6.18 | 0.01 | 0.31 | 0.76 |
| Age |  |  |  | 0.22 | 0.13 | 0.08 | 1.67 | 0.10 |
| Any other illness |  |  |  | 0.59 | 3.82 | 0.01 | 0.15 | 0.88 |
| PCS |  |  |  | 0.98 | 0.14 | 0.34 | 7.21 | 0.00 |
| **Step 3** | .42 | .177 | .06 |  |  |  |  |  |
| Sex |  |  |  | 5.81 | 6.06 | .04 | 0.96 | .34 |
| Age |  |  |  | 0.23 | 0.13 | .08 | 1.74 | .08 |
| Any other illness |  |  |  | -0.79 | 3.71 | -.01 | -0.21 | .83 |
| PCS |  |  |  | 0.78 | 0.14 | .27 | 5.63 | <.001 |
| Anxiety change |  |  |  | 0.07 | 0.05 | .09 | 1.37 | .17 |
| Mood change |  |  |  | 0.10 | 0.06 | .11 | 1.77 | .08 |
| Exercise change |  |  |  | 0.10 | 0.03 | .16 | 3.45 | .001 |

## 4. Discussion

We set out to understand how the COVID-19 pandemic, and associated lockdown restrictions, impacted individuals with chronic pain in terms of their psychological wellbeing, physical activity levels and pain experience. The findings reveal that people with chronic pain reported self-perceived increases in levels of pain severity during the most stringent period of lockdown in the UK (mid-April to early-May 2020) compared to the period before lockdown. They were more adversely affected by lockdown conditions than pain-free individuals, reporting greater self-perceived increases in anxiety and depressed mood, increased loneliness and reduced levels of physical exercise. People with chronic pain were more likely to be self-isolating due to high-risk status (observing increased levels of social-distancing and restrictions on activity) and more likely to report any other illness in the preceding fortnight compared to non-pain counterparts. We hypothesised a mediating role for pain catastrophizing on perceived changes in pain during lockdown and its mental and physical health consequences. The extent of self-perceived increases in pain symptoms in individuals with chronic pain was magnified by greater levels of pain catastrophizing, which also accounted for some of the impact of decreased mood on perception of pain. Perceived decreases in levels of physical exercise also independently predicted perceptions of increased pain. Interestingly, actual changes in pain severity (relative to pre-lockdown reports of pain measured in a subgroup with baseline data) did not change significantly. Yet, patients in this subgroup still reported self-perceived pain increases during lockdown, which were also predicted by baseline levels of pain catastrophizing. Overall, the findings suggest that during this period of crisis, pain catastrophizing and physical activity levels are important targets for pain-management interventions.

Pain catastrophizing and reduced levels of exercise are both are essential components of the fear-avoidance model of chronic pain [19; 29; 55]. In chronic pain populations, pain catastrophizing contributes to hypervigilance and fear related to pain and results in lower levels of psychological resilience [49]. People with high pain catastrophizing scores have been shown to avoid strenuous exercise [52]. Research evidence from chronic pain patients demonstrates that catastrophizing predicts psychological distress [48], avoidance of daily living activities and increased levels of physical dysfunction [56]. Physical inactivity promotes physical deconditioning, which then exacerbates pain during activity to cause greater aversion in a cycle of fear-avoidance [29; 54]. In this manner, pain catastrophizing promotes behavioural responses which lead to exacerbation of pain and other symptoms in chronic pain patients contributing to reduced quality of life [19].

In the present research, pain catastrophizing mediated the relationship between lockdown related changes in perceived pain and depressed mood which reflects a recent study of older chronic pain patients [13]. Pain catastrophizing was previously shown to mediate the relationship between negative interpersonal events and pain-related affective symptoms [50] and also moderated effects of exposure to missile attacks on pain and depressed mood in chronic pain patients in Israel [38]. Together, these studies highlight the importance of catastrophizing in chronic pain populations during the response to negative, high stress situations. No other studies have yet analysed levels of pain catastrophizing in the general population during the COVID-19 pandemic. However, health anxiety, which causes one to amplify perception of bodily sensations or changes as symptoms of being ill and which impacts on chronic pain experience [5], was recently shown to be exacerbated by the current pandemic [6], particularly in vulnerable populations [16].

It was recently highlighted that public health, social, clinical and psychological factors point to the likelihood of increased risk of pain and other symptoms in chronic pain populations during the COVID-19 pandemic [21]. The present findings confirm this risk, and demonstrate that people with chronic pain are more adversely affected by lockdown conditions compared to pain-free individuals. Perceived increases in pain severity and psychological distress offer empirical support to calls for rapid measures to provide appropriate care provision to chronic pain patients throughout this period [21; 47]. Cognitive behavioural therapy for chronic pain has greatest effectiveness when specifically targeting high catastrophizing patients [46], and can be successfully delivered using remote technology to reduce pain catastrophizing [9]. Remote technologies also have the potential to deliver pain physiology education, which are effective in alleviating pain catastrophizing [34; 36]. Physical exercise interventions also offer a flexible and potentially effective approach [3; 24]. Meta-analyses of telemedicine approaches for pain management provision and exercise therapy in chronic pain patients indicate positive outcomes that are broadly comparable to usual care [2; 40], and highlight that telemedicine options may be a suitable substitute when usual care is not possible. Based on our data, we contend that remote pain management approaches to reduce pain catastrophizing (particularly in high catastrophizing patients) and promote physical activity should be considered for rapid implementation during the current crisis.

The findings from the baseline group indicated that, although self-reported levels of pain severity are perceived to increase during lockdown, actual levels of pain reported are comparable to data recorded before the pandemic. There are a number of reasons why this may be the case. Firstly, it could indicate that self-perceived increases in pain severity are not due to actual increases in physical pain, but more a consequence of increased psychological distress. In the present study, baseline pain catastrophizing levels predicted the degree of self-perceived pain increase in the baseline subgroup. Previously, prospective studies have shown that baseline pain catastrophizing predict severity of post-operative pain [26; 45]. As pain catastrophizing was also the strongest predictor of self-perceived increases in pain in the full chronic pain cohort, this points to the need to make this a principle clinical outcome and target for telemedicine pain management. On the other hand, it must be noted that the baseline sample was selected from ongoing or previous research which utilised local pain clinics for recruitment. These respondents did exhibit some demographic differences (older, greater proportion of males), and significantly higher levels of pain severity, relative to chronic pain respondents recruited through other methods (Appendix 1). There could also be differences due to the data collection methods, with lockdown data collected using online tools compared to face-to-face clinics which could promote demand characteristics. Furthermore, actual changes in pain were measured using a different question and response scale compared to retrospective change ratings, pointing to the possibility that the latter method maybe more sensitive to measuring changes in pain, albeit less quantifiable in terms of actual pain severity. Overall, we caution that the finding of no actual pain increases, compared to pre-lockdown data, in the baseline subgroup should be interpreted with restraint.

The present research has some limitations. Firstly the chronic pain group were more adversely affected on *all* included measures of interest, although we could not practically include every clinically relevant measure available. For example, pain acceptance and perceived self-efficacy could be important factors not captured here. The relevant tools to capture these contributors typically include items that discuss lifestyle capability in the context of pain (e.g., ‘I lead a full life even though I have chronic pain’) in the Chronic Pain Acceptance Questionnaire [32]. There was a high risk that the validity of such items would be negatively impacted by the confounding effect of lockdown restrictions on lifestyle. The decision to focus on pain catastrophizing in the present study also reflects the fact that negative thought patterns have been shown to be more closely related to outcomes of perceived pain severity than positive factors such as pain acceptance [44]. Finally, as a cross sectional design, it is not possible to infer the causal nature of relationships between the many biopsychosocial factors which could be impacted during the current pandemic. In light of this, it is worth highlighting that this study reports on the first phase of data in a longitudinal design. Current respondents will continue to report on these measures regularly across coming months. Longitudinal data will permit more complex analyses to consider causality of the relationship between pain severity, pain cognition, psychological and physical wellbeing. It will also allow for consideration of longer-term impacts of lockdown restrictions which could yet have unforeseen and far reaching implications.

To conclude, the current findings are important because they represent the first empirical data to highlight increased suffering in people with chronic pain during lockdown. Specifically, people with chronic pain reported self-perceived increases in pain levels as well as increased adverse effects of lockdown compared to the non-pain comparison group. The findings support the urgent need for efforts to adapt remote clinical provision to mitigate unnecessary suffering. We also highlight the pivotal role of pain catastrophizing and reduced physical activity on the experience of people who live with chronic pain during lockdown conditions. This is significant because it points to potential clinical targets for therapeutic and behavioural interventions during the current, and future, crises. It may be possible to rapidly adapt efforts to rapidly target remote pain management toward reducing levels of pain catastrophizing, particularly in high catastrophizing patients, and promote physical activity as the pandemic continues. However, to truly establish whether such measures would be beneficial would require prospective research, preferably within a RCT design.

## Data Availability

The data that support the findings of this study will be openly available.

https://figshare.com/account/projects/81860/articles/12424661

## 5. Acknowledgements

We are grateful to Miss Rosalie Palmer for assistance with recruitment. Dr Fallon has no conflicts of interest to disclose, the rapid requirement for the research prevented pre-registration procedures. This study was supported by the Pain Relief Foundation, UK.

## Notes

### Competing Interest Statement

The authors have declared no competing interest.

### Clinical Trial

The research was rapidly established following implementation of lockdown restrictions in response to the COVID-19 pandemic.

### Author Declarations

The study was conducted in line with the recommendations of the Declaration of Helsinki and was approved by the local University of Liverpool Research Ethics Committee.

## References

[1] Guidance on social distancing for everyone in the UK Vol. 2020. https://www.gov.uk/government/publications/covid-19-guidance-on-social-distancing-andfor-vulnerable-people/guidance-on-social-distancing-for-everyone-in-the-uk-and-protectingolder-people-and-vulnerable-adults: Public Health England, 2020.

[2] Adamse C, Dekker-Van Weering MG, van Etten-Jamaludin FS, Stuiver MM. The effectiveness of exercise-based telemedicine on pain, physical activity and quality of life in the treatment of chronic pain: A systematic review. J Telemed Telecare 2018;24(8):511–526.

[3] Ambrose KR, Golightly YM. Physical exercise as non-pharmacological treatment of chronic pain: Why and when. BEST PRACT RES CL RH 2015;29(1):120–130.

[4] Armitage R, Nellums LB. COVID-19 and the consequences of isolating the elderly. The Lancet Public Health 2020;5(5):e256.

[5] Asmundson GJG, Katz J. Understanding the co-occurrence of anxiety disorders and chronic pain: state-of-the-art. Depress Anxiety 2009;26(10):888–901.

[6] Asmundson GJG, Taylor S. How health anxiety influences responses to viral outbreaks like COVID-19: What all decision-makers, health authorities, and health care professionals need to know. J Anxiety Disord 2020;71:102211.

[7] Bondesson E, Larrosa Pardo F, Stigmar K, Ringqvist Å, Petersson IF, Jöud A, Schelin MEC. Comorbidity between pain and mental illness – Evidence of a bidirectional relationship. Eur J Pain 2018;22(7):1304–1311.

[8] Breivik H, Eisenberg E, O’Brien T. The individual and societal burden of chronic pain in Europe: the case for strategic prioritisation and action to improve knowledge and availability of appropriate care. BMC Public Health 2013;13(1):1229.

[9] Buhrman M, Nilsson-Ihrfeldt E, Jannert M, Ström L, Andersson G. Guided internet-based cognitive behavioural treatment for chronic back pain reduces pain catastrophizing: a randomized controlled trial. Journal of rehabilitation medicine2011;43(6):500–505.

[10] Burckhardt CS, Clark SR, Bennett RM. The fibromyalgia impact questionnaire: development and validation. J Rheumatol 1991;18(5):728–733.

[11] Chang M-H, Hsu J-W, Huang K-L, Su T-P, Bai Y-M, Li C-T, Yang AC, Chang W-H, Chen T-J, Tsai S-J, Chen M-H. Bidirectional Association Between Depression and Fibromyalgia Syndrome: A Nationwide Longitudinal Study. J Pain 2015;16(9):895–902.

[12] Chen Y, Zhou H, Zhou Y, Zhou F. Prevalence of self-reported depression and anxiety among pediatric medical staff members during the COVID-19 outbreak in Guiyang, revalence of self-reported depression and anxiety among pediatric medical staff members during the COVID-19 outbreak in Guiyang China. Psychiatry Res2020;288:113005.

[13] Cheng ST, Leung CMC, Chan KL, Chen PP, Chow YF, Chung JWY, Law ACB, Lee JSW, Leung EMF, Tam CWC. The relationship of self-efficacy to catastrophizing and depressive symptoms in community-dwelling older adults with chronic pain: A moderated mediation model. PloS one2018;13(9):e0203964.

[14] Clauw DJ, Engel CC, Jr., Aronowitz R, Jones E, Kipen HM, Kroenke K, Ratzan S, Sharpe M, Wessely S. Unexplained Symptoms After Terrorism and War: An Expert Consensus Statement. J Occup Environ Med2003;45(10).

[15] Cleeland CS, Ryan KM. Pain assessment: Global use of the Brief Pain Inventory. Annals, ain assessment: Global use of the Brief Pain Inventory. Annals Academy of Medicine, Singapore1994;23(2):129–138.

[16] Corbett GA, Milne SJ, Hehir MP, Lindow SW, O’Connell M P. Health anxiety and behavioural changes of pregnant women during the COVID-19 pandemic. Eur J Obstet Gynecol Reprod Biol 2020.

[17] Coronaviridae Study Group of the International Committee on Taxonomy of V. The species Severe acute respiratory syndrome-related coronavirus: classifying 2019-nCoV and naming it SARS-CoV-2, Vol. 5, 2020. pp. 536–544.

[18] Cottam WJ, Condon L, Alshuft H, Reckziegel D, Auer DP. Associations of limbic-affective brain activity and severity of ongoing chronic arthritis pain are explained by trait anxiety. NeuroImage: Clinical 2016;12:269–276.

[19] Crombez G, Eccleston C, Van Damme S, Vlaeyen JW, Karoly P. Fear-avoidance model of chronic pain: the next generation. Clin J Pain 2012;28(6):475–483.

[20] Dominick CH, Blyth FM, Nicholas MK. Unpacking the burden: Understanding the relationships between chronic pain and comorbidity in the general population. PAIN 2012;153(2):293–304.

[21] Eccleston C, Blyth FM, Dear BF, Fisher EA, Keefe FJ, Lynch ME, Palermo TM, Reid MC, Williams ACdC. Managing patients with chronic pain during the COVID-19 outbreak: considerations for the rapid introduction of remotely supported (eHealth) pain management services. Pain 2020;161(5):889–893.

[22] Edwards RR, Cahalan C, Mensing G, Smith M, Haythornthwaite JA. Pain, catastrophizing, and depression in the rheumatic diseases. Nat Rev Rheumatol 2011;7(4):216–224.

[23] Fayaz A, Croft P, Langford RM, Donaldson LJ, Jones GT. Prevalence of chronic pain in the UK: a systematic review and meta-analysis of population studies. BMJ Open 2016;6(6):e010364.

[24] Geneen LJ, Moore RA, Clarke C, Martin D, Colvin LA, Smith BH. Physical activity and exercise for chronic pain in adults: an overview of Cochrane Reviews. Cochrane Database Syst Rev 2017(4).

[25] Giuseppe L, Brandon MH, Chiara B, Fabian S-G. Health risks and potential remedies during prolonged lockdowns for coronavirus disease 2019 (COVID-19). Diagnosis 2020;7(2):85–90.

[26] Granot M, Ferber SG. The Roles of Pain Catastrophizing and Anxiety in the Prediction of Postoperative Pain Intensity: A Prospective Study. Clin J Pain 2005;21(5).

[27] Jakobsson J, Malm C, Furberg M, Ekelund U, Svensson M. Physical Activity During the Coronavirus (COVID-19) Pandemic: Prevention of a Decline in Metabolic and Immunological Functions. Front Sports Act Living 2020;2(57).

[28] Kroenke K, Wu J, Bair MJ, Krebs EE, Damush TM, Tu W. Reciprocal relationship between pain and depression: a 12-month longitudinal analysis in primary care. J Pain 2011;12(9):964–973.

[29] Leeuw M, Goossens ME, Linton SJ, Crombez G, Boersma K, Vlaeyen JW. The fear-avoidance model of musculoskeletal pain: current state of scientific evidence. J Behav Med 2007;30(1):77–94.

[30] Li J, Yang Z, Qiu H, Wang Y, Jian L, Ji J, Li K. Anxiety and depression among general population in China at the peak of the COVID-19 epidemic. World Psychiatry 2020;19(2):249–250.

[31] Li X, Dai T, Wang H, Shi J, Yuan W, Li J, Chen L, Zhang T, Zhang S, Kong Y, Yue N, Shi H, He Y, Hu H, Liu F, Yang C. [Clinical analysis of suspected COVID-19 patients with anxiety and depression]. Zhejiang Da Xue Xue Bao Yi Xue Ban 2020;49(2):203–208.

[32] McCracken LM, Vowles KE, Eccleston C. Acceptance of chronic pain: component analysis and a revised assessment method. Pain 2004;107(1–2):159–166.

[33] McWilliams LA, Cox BJ, Enns MW. Mood and anxiety disorders associated with chronic pain: an examination in a nationally representative sample. Pain 2003;106(1):127–133.

[34] Meeus M, Nijs J, Van Oosterwijck J, Van Alsenoy V, Truijen S. Pain physiology education improves pain beliefs in patients with chronic fatigue syndrome compared with pacing and self-management education: a double-blind randomized controlled trial. Archives of physical medicine and rehabilitation2010;91(8):1153–1159.

[35] Moric M, Buvanendran A, Lubenow TR, Mehta A, Kroin JS, Tuman KJ. Response of Chronic Pain Patients to Terrorism: The Role of Underlying Depression. Pain Med 2007;8(5):425–432.

[36] Moseley GL, Nicholas MK, Hodges PW. A randomized controlled trial of intensive neurophysiology education in chronic low back pain. Clin J Pain 2004;20(5):324–330.

[37] Newman MG, Zainal NH. The value of maintaining social connections for mental health in older people. Lancet Public Health 2020;5(1):e12-e13.

[38] Noyman-Veksler G, Shalev H, Brill S, Rudich Z, Shahar G. Chronic pain under missile attacks: Role of pain catastrophizing, media, and stress-related exposure. Psychol Trauma 2018;10(4):463–469.

[39] Ozamiz-Etxebarria N, Dosil-Santamaria M, Picaza-Gorrochategui M, Idoiaga-Mondragon N. Stress, anxiety, and depression levels in the initial stage of the COVID-19 outbreak in a population sample in the northern Spain. Cadernos de Saude Publica 2020;36(4):e00054020.

[40] Piga M, Cangemi I, Mathieu A, Cauli A. Telemedicine for patients with rheumatic diseases: Systematic review and proposal for research agenda. Seminars in Arthritis and Rheumatism 2017;47(1):121–128.

[41] Ploghaus A, Narain C, Beckmann CF, Clare S, Bantick S, Wise R, Matthews PM, Rawlins JNP, Tracey I. Exacerbation of Pain by Anxiety Is Associated with Activity in a Hippocampal Network. The Journal of Neuroscience 2001;21(24):9896.

[42] Reger MA, Stanley IH, Joiner TE. Suicide Mortality and Coronavirus Disease 2019—A Perfect Storm? JAMA Psychiatry 2020.

[43] Rice ASC, Smith BH, Blyth FM. Pain and the global burden of disease. PAIN 2016;157(4).

[44] Richardson EJ, Ness TJ, Doleys DM, Baños JH, Cianfrini L, Scott Richards J. Depressive symptoms and pain evaluations among persons with chronic pain: Catastrophizing, but not pain acceptance, shows significant effects. Pain2009;147(1):147–152.

[45] Riddle DL, Wade JB, Jiranek WA, Kong X. Preoperative Pain Catastrophizing Predicts Pain Outcome after Knee Arthroplasty. CLIN ORTHOP RELAT R 2010;468(3):798–806.

[46] Schütze R, Rees C, Smith A, Slater H, Campbell JM, O’Sullivan P. How Can We Best Reduce Pain Catastrophizing in Adults With Chronic Noncancer Pain? A Systematic Review and Meta-Analysis. J Pain 2018;19(3):233–256.

[47] Shanthanna H, Strand NH, Provenzano DA, Lobo CA, Eldabe S, Bhatia A, Wegener J, Curtis K, Cohen SP, Narouze S. Caring for patients with pain during the COVID-19 pandemic: consensus recommendations from an international expert panel. Anaesthesia 2020;n/a (n/a).

[48] Sturgeon JA, Zautra AJ. State and Trait Pain Catastrophizing and Emotional Health in Rheumatoid Arthritis. Ann Behav Med 2012;45(1):69–77.

[49] Sturgeon JA, Zautra AJ. Psychological Resilience, sychological Resilience Pain Catastrophizing, and Positive Emotions: Perspectives on Comprehensive Modeling of Individual Pain Adaptation. Curr Pain Headache Rep2013;17(3):317.

[50] Sturgeon JA, Zautra AJ, Arewasikporn A. A multilevel structural equation modeling analysis of vulnerabilities and resilience resources influencing affective adaptation to chronic pain. Pain 2014;155(2):292–298.

[51] Sullivan MJL, Bishop SR, Pivik J. The Pain Catastrophizing Scale: Development and validation. Psych Ass 1995;7:524–532.

[52] Sullivan MJL, Rodgers WM, Wilson PM, Bell GJ, Murray TC, Fraser SN. An experimental investigation of the relation between catastrophizing and activity intolerance. Pain 2002;100(1):47–53.

[53] Tull MT, Edmonds KA, Scamaldo KM, Richmond JR, Rose JP, Gratz KL. Psychological Outcomes Associated with Stay-at-Home Orders and the Perceived Impact of COVID-19 on Daily Life. Psychiatry research 2020;289:113098–113098.

[54] Turk DC, Wilson HD. Fear of Pain as a Prognostic Factor in Chronic Pain: Conceptual Models, ear of Pain as a Prognostic Factor in Chronic Pain: Conceptual Models Assessment, and Treatment Implications. Curr Pain Headache Rep2010;14(2):88–95.

[55] Vlaeyen JWS, Linton SJ. Fear-avoidance and its consequences in chronic musculoskeletal pain: a state of the art. Pain 2000;85(3):317–332.

[56] Westman AE, Boersma K, Leppert J, Linton SJ. Fear-avoidance beliefs, catastrophizing, and distress: a longitudinal subgroup analysis on patients with musculoskeletal pain. Clin J Pain 2011;27(7):567–577.

[57] Zhuo M. Neural Mechanisms Underlying Anxiety–Chronic Pain Interactions. Trends Neurosci 2016;39(3):136–145.

[58] Zigmond AS, Snaith RP. The hospital anxiety and depression scale. Acta psychiatrica Scandinavica 1983;67(6):361–370.

